# Tracking COVID-19 in England and Wales: Insights from Virus Watch - a prospective community cohort study

**DOI:** 10.1101/2023.12.19.23299951

**Authors:** Wing Lam Erica Fong, Vincent Grigori Nguyen, Sarah Beale, Thomas E Byrne, Cyril Geismar, Ellen Fragaszy, Jana Kovar, Annalan M D Navaratnam, Alexei Yavlinsky, Ibrahim Abubakar, Andrew C Hayward, Robert W Aldridge

## Abstract

**Background:** Virus Watch is a prospective community cohort study of COVID-19 of 28□,527 households in England and Wales designed to estimate the incidence of PCR-confirmed COVID-19 in those with respiratory presentations and examine symptom profiles and transmission of COVID-19 in relation to population movement and behaviour. The Office for National Statistics (ONS) COVID-19 infection survey (CIS) was the largest regular survey of COVID-19 infections and antibodies in the UK and included 227,797 households. In this analysis, we aimed to compare incidence rate estimates from the two studies to understand differences in estimates from the two study designs.

**Methods:** We used the Virus Watch prospective community cohort study to estimate the overall SARS-CoV-2 incidence rate and incidence rate by age in England and Wales from June 2020 to February 2023. Virus Watch data consisted of self-reported laboratory COVID-19 test results and linkage to the Second Generation Surveillance System, the UK national database for COVID-19 testing. We compared our findings with modelled incidence rates from ONS CIS using 3-day rolling Pearson’s correlation to measure synchrony.

**Results:** 58,628 participants were recruited into the Virus Watch study between June 2020 and March 2022, of whom 52,526 (90%) were reported to be living in England and 1,532 (2.6%) in Wales. COVID-19 incidence rates were initially similar across age groups until the Delta wave when rates increased at different magnitudes. During the Omicron BA.1, the 0-14 age group had the highest incidence rates, which shifted to the 25-44 age group with Omicron BA.2, 4, and 5 dominance. We found strong synchrony between Virus Watch and ONS CIS COVID-19 incidence estimates for England and Wales, both with and without the incorporation of linked national testing data into the Virus Watch study. In particular, the magnitude and trend of Virus Watch- and ONS-estimated rates for England were generally consistent, although Virus Watch-estimated peaks of infection during the Omicron BA.1 and 2 waves were found to be lower than estimates from the ONS.

**Conclusion:** Our findings suggest that the Virus Watch research approach is a low-cost and effective method for on-going surveillance of COVID-19 regardless of the availability of national testing in the UK. Similar approaches can also be utilised by low-resource settings to provide accurate incidence rate estimates to better monitor and respond to COVID-19 as well as other acute respiratory diseases in the future.

## Introduction

Since the global outbreak of Severe acute respiratory syndrome coronavirus 2 (SARS-CoV-2), numerous large-scale studies such as the Office for National Statistics (ONS) COVID-19 infection survey (CIS) and the REal-time Assessment of Community Transmission-1 (REACT-1) Study, have been established in the United Kingdom (UK) to monitor COVID-19 transmission [1, 2]. These studies have been used to estimate the incidence and prevalence of SARS-CoV-2 infections and subsequent burden of COVID-19. Such research has been crucial for monitoring infection rates, thus enabling the timely implementation of policies and interventions to mitigate the spread of COVID-19.

Conducting large-scale epidemiological studies to accurately estimate infection rate trends can be a resource-intensive and costly process that necessitates sufficient testing coverage and significant testing capacity [3, 4]. The ONS CIS is a large longitudinal study which began on 26 April 2020. It consists of a randomly selected, representative sample of children and adults (aged 2 and over) in private residential households (excluding care homes and other communal establishment settings) [1]. Nose and throat swabs were obtained from study participants on a weekly basis regardless of symptoms and tested for SARS-CoV-2 using reverse transcriptase polymerase chain reaction (RT-PCR), which identifies active infections at the time of sample collection. By September 2020, around 150,000 polymerase chain reaction (PCR) tests were conducted per month, and the study further expanded to analyse an average of around 390,300 PCR results each month between May 2021 and March 2022. The ONS CIS incentivised the return of swabs and as of 18th November 2021 the total number of vouchers issued was 6,918,402, with a face value of £211,522,225 [5]. From 2020 to 2023, ONS had directly attributed a total of £988.5 million in costs to the study [6]. Such a substantial financial investment may often be unattainable in developing countries or regions, as well as high income settings in the absence of a pandemic, and therefore, limit their capacity to conduct and maintain effective disease surveillance [3]. This underscores the importance of exploring cost-effective research approaches to ensure that such countries and regions can track the spread of COVID-19 as well as other acute respiratory diseases without incurring insurmountable costs.

Virus Watch study is a community cohort study conducted in England and Wales with a spend of £4.89 million. Study data primarily consists of online self-reported laboratory COVID-19 test results and linkage to the Second Generation Surveillance System (SGSS), the UK national database for COVID-19 testing. These test results were likely from free PCR or lateral flow tests (LFTs) that became widely available through the national Test Trace and Isolate Programme which began in May 2020 [7]. Individuals experiencing a high temperature, new, continuous cough or loss or change to your sense of smell or taste were recommended to undergo testing [8]. Furthermore, since Virus Watch participants were not incentivised to take nose and throat swabs, the majority of the testing conducted was symptomatic.

## Objectives

Virus Watch and the ONS CIS had key differences in study design including recruitment strategy (non-random sampling vs. random sampling), incentivisation for return of swabs (none vs. incentives) and indications for testing (symptomatic based on national guidelines vs. regular asymptomatic testing). In this study, we aimed to compare modelled incidence rate estimates from June 2020 to February 2023 from Virus Watch to ONS CIS in order to assess the validity of Virus Watch results in capturing infection rates given differences in our study design.

## Methods

### Study design and participants

Virus Watch is a large prospective household cohort study of the transmission and burden of COVID-19 in England and Wales. The study began on 24th June 2020 and is ongoing as of 20 October 2023. Between June 2020 and March 2022, a total of 28,527 households and 58,628 participants aged 0-98 years (mean age: 48) were recruited [9]. The full study design and methodology has been described elsewhere [9, 10], with relevant elements for the present study outlined here.

### Procedure

We triangulated our estimated COVID-19 incidence rates with those from the ONS CIS. In Virus Watch, we used multiple sources to identify SARS-CoV-2 infections among study participants. Infection was identified based on the first positive results from the following sources:

1. Data linked to the SGSS, which contains SARS-CoV-2 test results using data from hospitalisations (Pillar 1), and community testing (Pillar 2). Linkage was conducted by NHS Digital with the linkage variables being sent in March 2021.The linkage period for SGSS Pillar 1 encompassed data from March 2020 until August 2021 and from June 2020 until November 2021 for Pillar 2. Linked data was only available for participants who registered with an English postcode.
2. Self-reported SARS-CoV-2 test results (PCR or LFT) received from outside the study (e.g. via the UK Test Trace and Isolate system or privately obtained) as part of the weekly illness survey.
3. Nasopharyngeal swab samples for PCR assays for SARS-CoV-2 collected from a subset of participants between October 2020 and May 2021 (n = 12,877). Participants carried out and posted a self-administered PCR swab if they experienced any of the following symptoms: fever, cough or loss or change of taste or smell for two or more days. All PCR swabs were tested for SARS-CoV-2 ribonucleic acid (RNA) via RT-PCR.
4. Nasopharyngeal swab samples for PCR assays for SARS-CoV-2 collected from a subset of participants starting in January 2023 (n = 2,851). Swab collection ended in June 2023. A PCR swab is self-administered if participants experience any of the following symptoms: fever, cough or loss or change of taste or smell and/or if they obtain a positive LFT result. All PCR swabs are tested for SARS-CoV-2 RNA via RT-PCR.

For each week, we identified the total number of participants who had or reported a positive PCR swab or LFT for SARS-CoV-2 at any point during the week and the total number of participants at-risk of SARS-CoV-2 infection during the week. These were used to calculate the 7-day average incidence rate per 10,000 people using the following equation:

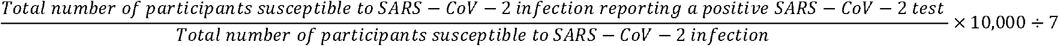

The denominator is composed of participants who did not test positive for SARS-CoV-2 in the past 90 days from the final day (Sunday) of each week. Participants included were those who completed the weekly survey reporting no illness or a negative SARS-CoV-2 test result, as well as those who had a negative SARS-CoV-2 test result from linked SGSS/Pillar 2 data for that particular week.

The ONS CIS calculated the incidence of COVID-19 from the Bayesian multilevel regression and poststratification model of positivity. Biased positivity estimates due to non-participation were corrected for potential non-representativeness by post-stratifying for age, sex, and region. The full details of ONS CIS study and methodology has been described elsewhere [11, 12]. The official reported estimates of incidence of COVID-19 of England and Wales are publicly available and obtained from /web/20231102114215/https://www.ons.gov.uk/peoplepopulationandcommunity/healthandsocialcare/conditionsanddiseases/datasets/coronaviruscovid19infectionsurveydata. COVID-19 incidence rate estimates per 10,000 people per day were provided between 8 June 2020 and 14 June 2022 and between 20 December 2022 and 14 February 2023.

We first plotted Virus Watch-estimated COVID-19 incidence rates over time by age group (0-14, 15-24, 25-44, 45-64, 65+). Subsequently, we calculated incidence rates among Virus Watch participants aged two and over and plotted it with ONS-estimated COVID-19 incidence rates from 26 April 2020 to 14 February 2022 - the final date for which ONS-estimated rates were available. We then used 3-day rolling Pearson’s correlation to measure synchrony between the two datasets. A histogram was plotted to show the frequency distribution of the correlation coefficients.

In addition, we estimated the total number of people who have been infected with COVID during different periods of time when different COVID-19 variants were most common. A variant was considered dominant if an estimated 60% or greater of COVID-19 infections were of that variant at the national level [13, 14]. These periods include:

1. Wild type: 26 April 2020 to 7 December 2020
2. Alpha: 8 December 2020 to 17 May 2021
3. Delta: 18 May 2021 to 13 December 2021
4. Omicron BA.1: 14 December 2021 to 21 February 2022
5. Omicron BA.2: 22 February 2022 to 6 June 2022
6. Omicron BA.4/5: 7 June 2022 to 11 November 2022

For this, we only included participants aged two and older with linked data (SGSS and Pillar 2) to minimise bias from loss-to-follow-up. The cumulative incidence by periods of time by variant of COVID-19 that was most common in England and cumulative incidence by variant period and age group (2-11, 12-16, 17-24, 25-34, 35-49, 50-69, 70+) were calculated. We estimated the number of people who tested positive for the first time during each specified period on any given day and aggregated this by period. The denominator for each time period is composed of participants susceptible to SARS-CoV-2 infection at the start of each period. Participants who were infected less than 90 days before or died before the start of/during each period were excluded. Virus Watch and ONS-estimated cumulative incidence by period and age group were then plotted.

## Results

**Table 1.** Demographics of Virus Watch study participants at baseline recruitment compared to the ONS CIS and 2021 Census.

| Characteristic | Virus Watch Participants<br>N (%) | ONS CIS<br>N (%) | Population of England<br>from ONS (%) |
| --- | --- | --- | --- |
| **All** | **58,628** | **535,121** | **-** |
| **Age group (years)\*** |  |  |  |
| 0-14 | 6,823 (12%) | - | 17% |
| 15-24 | 4,047 (6.9%) | - | 12% |
| 25-44 | 11,725 (20%) | - | 26% |
| 45-64 | 19,657 (34%) | - | 26% |
| 65+ | 16,376 (28%) | - | 19% |
| **Age group (years)\*** |  |  |  |
| 2-11 | 4,617 (8%) | 33,507 (6%) | 13% |
| 12-16 | 2,623 (5%) | 29,147 (5%) | 6% |
| 17-24 | 3,000 (5%) | 35,403 (7%) | 9% |
| 25-34 | 4,973 (9%) | 52,359 (10%) | 14% |
| 35-49 | 10,406 (18%) | 103,068 (19%) | 20% |
| 50-69 | 22,789 (39%) | 176,814 (33%) | 25% |
| 70+ | 9,589 (17%) | 104,823 (20%) | 14% |
| **Sex (including derived)\*\*** |  |  |  |
| Male | 26,274 (45%) | 252,047 (47%) | 49% |
| Female | 31,533 (54%) | 283,074 (53%) | 51% |
| Other/ Missing/ Prefer not to say | 821 (1.4%) | - | - |
| **Ethnicity** |  |  |  |
| White | 43,968 (75%) | 493,026 (92%) | 86% |
| Asian/ Asian British | 3,155 (5.4%) | 22,128 (4%) | 7% |
| Black/ African/<br>Caribbean/ Black<br>British | 493 (0.8%) | 5,476 (1%) | 3% |
| Mixed/ Multiple ethnic<br>groups | 998 (1.7%) | 9,598 (2%) | 2% |
| Other ethnic groups | 288 (0.5%) | 4,804 (1%) | 2% |
| Missing | 9,726 (17%) | 89 (0.01%) | - |
| **Region** |  |  |  |
| North East | 2,528 (4.3%) | - | 5% |
| North West | 5,572 (9.5%) | - | 12% |
| Yorkshire and The<br>Humber | 3,035 (5.2%) | - | 9% |
| East Midlands | 4,945 (8.4%) | - | 8% |
| West Midlands | 3,020 (5.2%) | - | 10% |
| East of England | 10,545 (18%) | - | 11% |
| London | 9,083 (15%) | - | 15% |
| South East | 9,845 (17%) | - | 15% |
| South West | 3,956 (6.7%) | - | 10% |
| Wales | 1,532 (2.6%) | - | 5% |
| Missing | 4,567 (7.8%) | - | - |

ONS CIS data obtained from *Coronavirus (COVID-19) Infection Survey: quality and methodology information (QMI)* [1]. England population estimates from ONS data for age, sex and region drawn from Population and household estimates, England and Wales: Census 2021, while data for ethnicity was drawn from Ethnic group, England and Wales: Census 2021.
\*Different categorisation of age groups were used in Virus Watch and ONS CIS.
\*\*Sex at birth was self-reported. If missing, sex was obtained via data linkage or derived via name-gender matching based on US names from 1930-2015 https://data.world/howarder/gender-by-name

From June 2020 to February 2023, a total of 58,628 participants were recruited into the Virus Watch study, 52,526 (90%) reported to be living in England and 1,532 (2.6%) in Wales. Compared with the population of England and Wales, the Virus Watch cohort is older, with a greater proportion of people in the 45-64 and 65+ age groups, and generally representative of both males and females. There is a lack of representativeness in some regions, including North West, South West, West Midlands and Yorkshire and The Humber, while regions such as East of England and South East were overrepresented. Some ethnic groups are also under-represented, notably the Black and Other Asian groups.

A total of 28,463, 92 and 29,609 positive PCR/LFT results were identified through self-report, Virus Watch swabs, and data linkage, respectively. Test results from the three sources were not mutually exclusive. From these results, we identified 30,031 COVID-19 cases in England and 570 in Wales between June 2020 and February 2023.

Using Virus Watch data, we estimated COVID-19 incidence rates by age group and found differing incidence rates throughout the course of the pandemic between the groups. During the periods when the wild type and Alpha variants were dominant, incidence rates for all age groups were similar, except at the beginning of the Alpha period where rates were higher for those aged 15-24 and 25-44. The rate for 0-14 year olds began to increase in September 2021 and eventually peaked at 259.6 cases per 10,000 per day (95% confidence interval (C.I.): 225.1, 297.9) during the Omicron BA.1 wave. Specifically during this period, we observed elevated COVID-19 incidence rates in the younger age groups (0-14, 15-24 and 25-44) compared to the 45-64 and 65+ age groups. With the emergence of Omicron BA.2, the highest incidence rate was observed among the 25-44 year old individuals particularly between March and April 2022. Concurrently, the highest incidence rates for the 45-64 and 65+ age groups were also recorded during this period. We estimated a rise in incidence rate as Omicron BA.4 and 5 became dominant, primarily within the 25-44 age group. By August 2022, the incidence rates across all age groups had decreased and were similar, although the rate for 25-44 and 45-64 age groups consistently remained higher in comparison to the other groups.

When comparing Virus Watch- and ONS-estimated incidence rates, the trend of Virus Watch-estimated incidence rate estimates of England correlated with that of the ONS across all waves of infection (Figure 2A). The magnitude of both rates were generally consistent, although Virus Watch-estimated peaks of infection during the Omicron BA.1 and 2 waves were found to be lower than estimates from the ONS. The overall Pearson estimate was 0.98 (p<0.01) which indicates a high global synchrony between ONS and Virus Watch estimated incidence rates in England over time. Furthermore, we observed a large number of coefficients close to +1 which suggests high local synchrony between the two estimates (Supplementary Figure 1). Without the inclusion of linked SGSS and Pillar 2 data, the Virus Watch-estimated incidence rates were consistently lower than that of ONS (Figure 2B). However, we continued to find high global (overall Pearson estimate: 0.96; p< 0.01) and local synchrony between the two estimates which indicates similar trends over time (Supplementary Figure 2).

**Figure 1.**
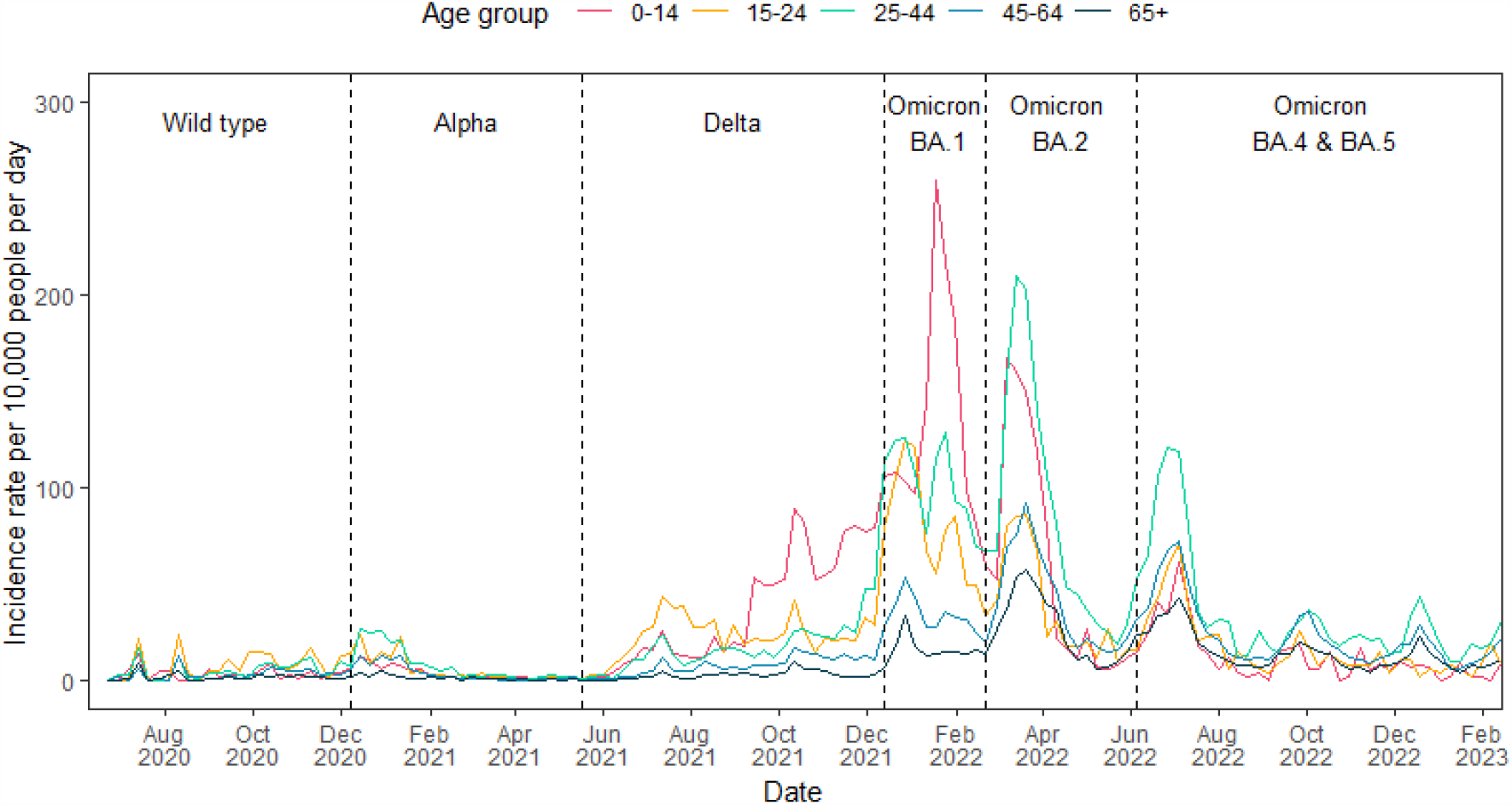
Virus Watch estimates of COVID-19 incidence rates in England by age group from June 2020 to February 2023 with the dominant variant of concern of each period labelled.

**Figure 2.**
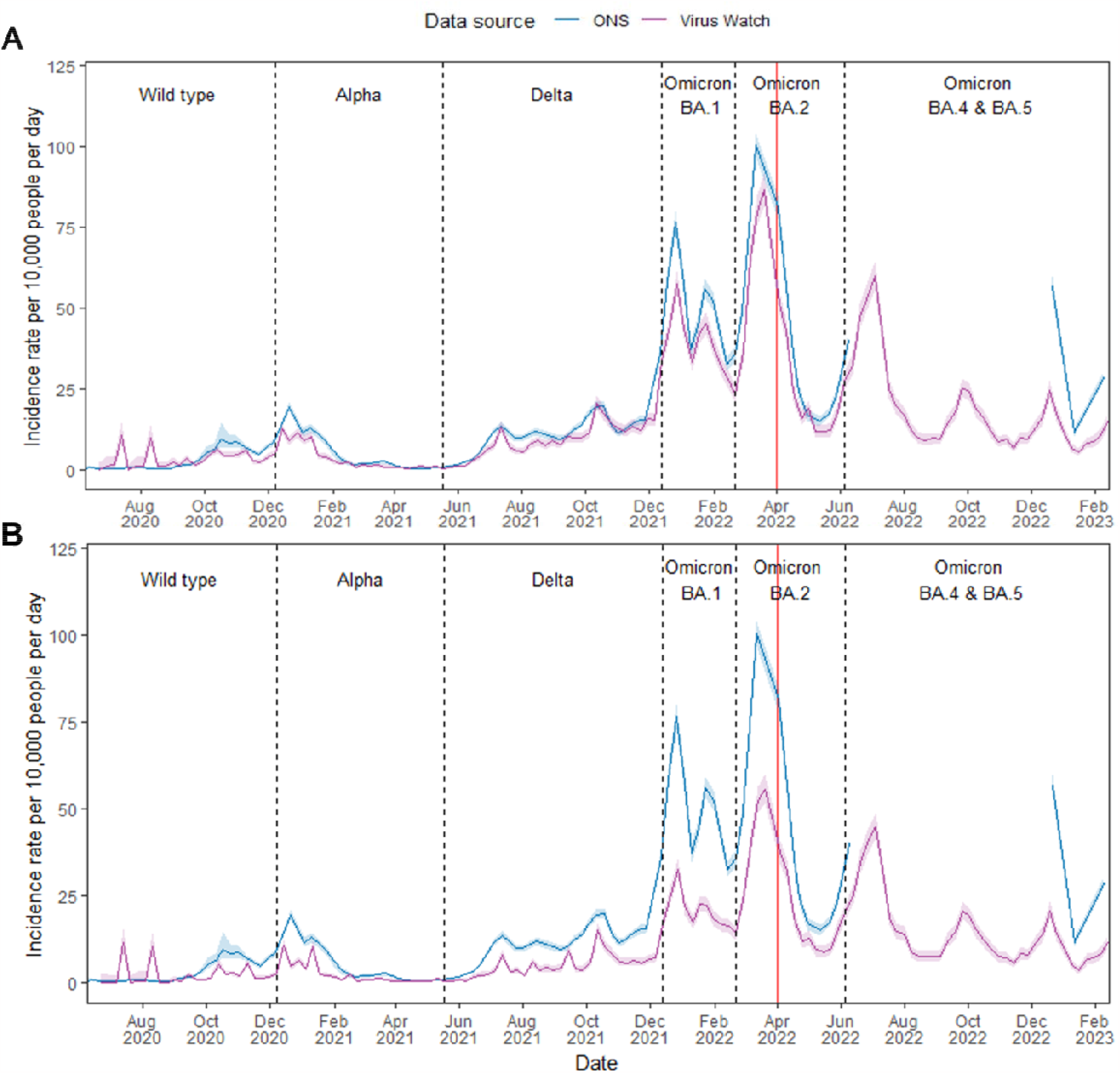
ONS CIS and Virus Watch estimates of COVID-19 incidence rates in England from June 2020 to February 2023 with the dominant variant of concern of each period labelled. Virus Watch data in A) incorporates SGSS and Pillar 2 linked data, while B) excludes SGSS and Pillar 2 linked data. Shaded regions indicate the 95% confidence interval for incidence rate estimates. Dotted vertical lines indicate the periods of time when different COVID-19 variants were most common in the UK, and the red line indicates the ending of national free COVID-19 testing.

In Wales, the overall trend of Virus Watch-estimated incidence rates over time was also consistent with ONS estimates (Figure 3). However, Virus Watch estimates exhibited greater fluctuations and large confidence intervals. The overall Pearson r was 0.78 (p <0.01) which indicated a high global synchrony between Virus Watch- and ONS-estimated incidence rates in Wales over time. When compared to the Virus Watch England incidence rate estimates, estimates for Wales have a lower synchrony evident by a fewer number of coefficients close to +1 (Supplementary Figure 3).

**Figure 3.**
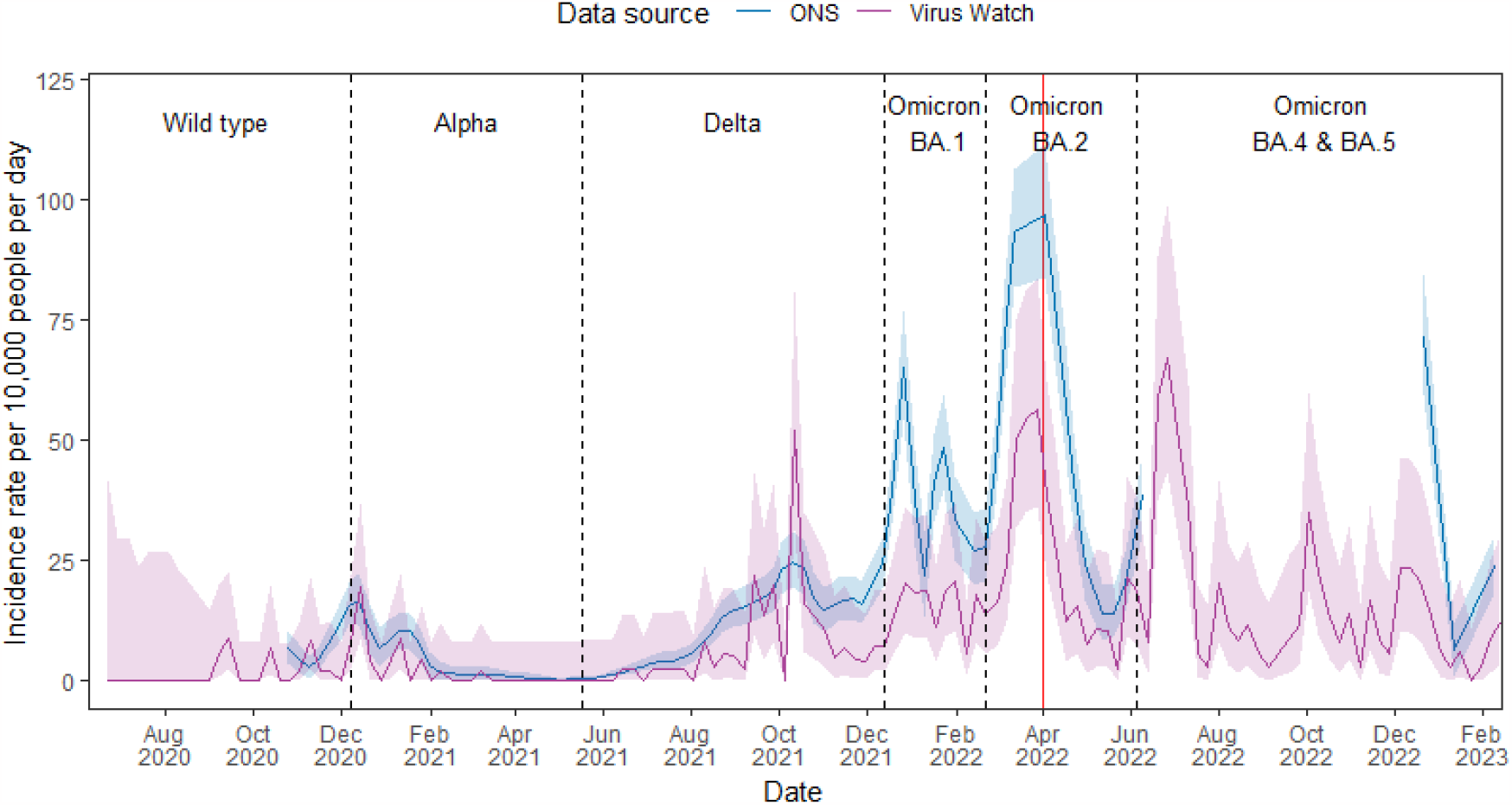
Virus Watch and ONS estimates of COVID-19 incidence rates in Wales from June 2020 to February 2023 with the dominant variant of concern of each period labelled. Shaded regions indicate the 95% confidence interval for incidence rate estimates. Dotted vertical lines indicate the periods of time when different COVID-19 variants were most common in the UK, and the red line indicates the ending of national free COVID-19 testing.

The trend of Virus Watch-estimated cumulative incidences by dominant variant periods in England were similar to that estimated by ONS but of lower magnitude (Supplementary Figure 4). Similarly, Virus Watch-estimated cumulative incidence by age group were also lower than ONS-estimated rates (Supplementary Figure 5). Virus Watch estimates captured an increase in cumulative incidence over time albeit not of the same magnitude when compared to that modelled by ONS among the younger populations (2-11, 12-16, 17-24, 25-34, 35-49) particularly during the Omicron BA.2 and BA.4 & 5 periods. This could be due to the end of free national testing that occurred during the Omicron BA.2 period.

## Discussion

In this analysis, we estimated COVID-19 incidence rates in England and Wales using Virus Watch data and triangulated them with ONS-estimates. Our findings demonstrate synchrony between Virus Watch and ONS incidence rates over time, particularly for England when linked national testing data in Virus Watch was incorporated. Without linked data, Virus Watch yielded lower incidence rate and cumulative incidence estimates than ONS, and this can be attributed to differing study designs. The ONS CIS study design employed a regular PCR swabbing protocol with participant incentives to ensure continued participation, enabling the detection of asymptomatic cases [1]. In contrast, Virus Watch primarily captured symptomatic cases through self-testing and reporting, with incentives provided only to recruits during a specific round of recruitment targeted at black, Asian and minority ethnic groups and no incentives were used to improve continued engagement [9]. Virus Watch-estimated incidence rates for Wales exhibited significant fluctuations over time, accompanied by large confidence intervals. This variability is likely a consequence of the small, non-representative Welsh cohort and the absence of linked national testing data in the Virus Watch study.

Ongoing COVID-19 surveillance is essential for early outbreak detection and identification of at-risk populations in order to facilitate implementation of effective control measures. It is also crucial for monitoring disease transmission and providing data for estimating hospitalisation and fatality rates associated with different variants of concern [15]. While the ONS CIS can be considered the ‘gold standard’ approach for surveillance, its application may pose challenges in low-income settings due to elevated costs, and could be unsustainable to maintain over extended durations in developed countries. Alternatively, surveillance utilising only national testing data is also valuable due to its timeliness and extensive coverage. However, compared to Virus Watch which collected extensive behavioural and medical risk factor information, the limited information collected through routine national surveillance would also impede our ability to identify at-risk populations and assess risk factors of infection [16]. Moreover, the viability of this surveillance method is also contingent upon the existence of comprehensive national testing infrastructure.

Our research approach was characterised by a strategic and cost-effective utilisation of resources for the estimation of COVID-19 incidence over time. This involved aggregating data derived from weekly surveys, invited PCR and LFT swab tests, and linkage to existing national testing programme datasets, specifically SGSS and Pillar 2 data. The incorporation of linked data not only minimised costs but also provided valuable insights into the dynamics of COVID-19 incidence throughout the pandemic. It also improved the reliability of the results by reducing the risk of underestimating COVID-19 incidence associated with the self-reporting of PCR/LFT test results (Figure 2A and 2B). In comparison to the ONS CIS and the national testing programme, our approach allowed the collection and analysis of multiple clinical risk factors and studying of symptom profiles associated with different SARS-CoV-2 variants of concern [17-26].

During periods of national testing, the Virus Watch approach may serve as a low-cost and supplementary surveillance method, while also allowing the consideration of multiple risk factors. In the absence of a comprehensive national testing programme, our study approach, primarily involving symptomatic self-swabbing, could yield acceptably accurate results with considerably lower costs than larger studies where swabbing is conducted irrespective of symptoms. This is evident from the Flu Watch study, which focused on influenza and employed a research approach similar to that of Virus Watch. The study demonstrated the effectiveness of this method in the absence of a national testing scheme, reporting an average 22-times higher (95% CI 17-28) rate of PCR-confirmed influenza across all winter seasons (2009-2011) than rates of PCR-confirmed disease from primary-care-based surveillance [27]. Our approach may also serve as a model for future acute respiratory disease surveillance in lower-resource settings, where limited funding and resources are common challenges.

## Limitations

Several important limitations should be considered in interpreting these findings. Due to the different study designs, we were unable to apply the identical methodology employed by ONS CIS to estimate COVID-19 incidence rates. While this suggests that our estimates are not directly comparable, observing similar estimates between the two studies is encouraging. Moreover, Virus Watch incidence rate and cumulative incidence estimates are likely to be underestimated as we predominantly captured only symptomatic cases and did not incentivise swabs to be sent back, in contrast to ONS CIS. It is also important to note that the smaller sample size in our study contributes to the limitations in the generalisability of our findings particularly for the lower age groups.

Despite being self-selected, the Virus Watch cohort is broadly representative of the general English population. However, the ONS CIS cohort has a higher level of representativeness due to their random sampling strategy [1]. As households were self-selected into the study, the Virus Watch cohort is biased towards participants with an interest in COVID-19 and health research, typically representing more cautious and health seeking individuals. Furthermore, the Virus Watch cohort skewed toward an older White British demographic, which limited our capacity to capture infections among ethnic minority groups, notably Black and Other Asian groups, and younger populations. Households with more than six members were not eligible for the study due to the limitations of the REDCap survey infrastructure, and people living in institutional settings such as care homes, university halls of residence and boarding schools were not eligible to participate, limiting the generalisability of findings for these groups. Additionally, Virus Watch participant retention rate decreased substantially to less than 50% over the three-year course of the study. This decline can be attributed to the relaxation of pandemic-related restrictions and loss of public interest in the pandemic. Notably, participants who have disengaged are typically younger, from an ethnic minority background and reside in London [9]. However, this level of attrition and demographic pattern is in line with other longitudinal cohort studies [28].

## Conclusion

We have presented a means of online weekly symptom and test reporting from over 58,000 participants to estimate the incidence of COVID-19 in England and Wales. By combining self-reported test results, invited swab tests and national testing programme data, we were able to obtain similar estimates to ONS CIS, a large-scale epidemiological study. We believe that our approach may serve as an cost-effective and supplementary method to track the spread of COVID-19 to guide public health response in the UK. Similarly, it can be implemented at a relatively low cost in both developed settings when routine testing and notification is not conducted and in low-resource settings. This will facilitate monitoring and response capabilities not only for COVID-19 but also other acute respiratory infectious diseases in the future.

## Acknowledgments

Ethics and Consent

The Virus Watch study was approved by the Hampstead NHS Health Research Authority Ethics Committee: 20/HRA/2320, and conformed to the ethical standards set out in the Declaration of Helsinki. All participants provided informed consent for all aspects of the study. Approval was also obtained from the Independent Group Advising on the Release of Data (IGARD) to use the NHS Digital data under DSA DARS-NIC-372269-N8D7Z-v1.6

## Data availability statement

Data from the Virus Watch cohort are available on ONS Secure Research Service. The data are available under restricted access as they contain sensitive health data. Access can be obtained by ONS Secure Research Service.

