## Supplementary materials for "Tracking COVID-19 in England and Wales: Insights from Virus Watch - a prospective community cohort study"

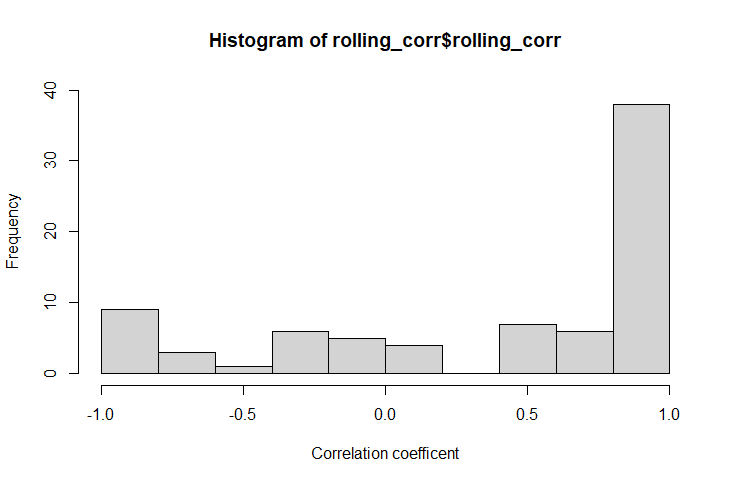

**Figure 1. Histogram of the frequency distribution of 3-day rolling Pearson correlation coefficients estimated using ONS and Virus Watch (including SGSS and Pillar 2 linked data) estimated COVID-19 incidence rates in England from 22 June 2020 to 13 June 2022.**

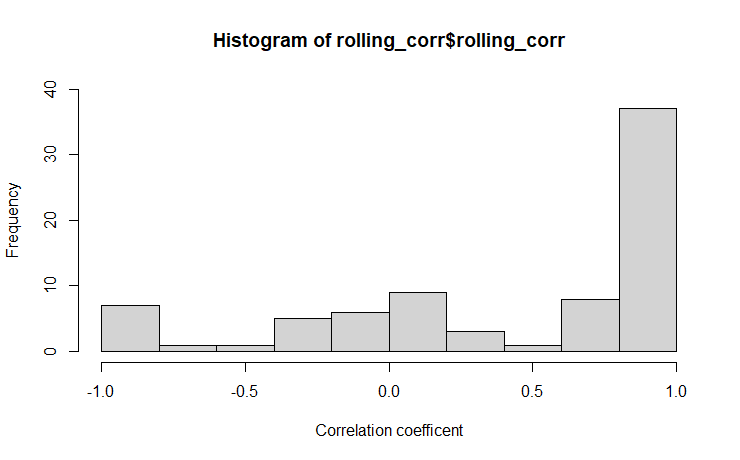

**Figure 2. Histogram of the frequency distribution of 3-day rolling Pearson correlation coefficients estimated using ONS and Virus Watch (excluding SGSS and Pillar 2 linked data) estimated COVID-19 incidence rates in England from 22 June 2020 to 13 June 2022.**

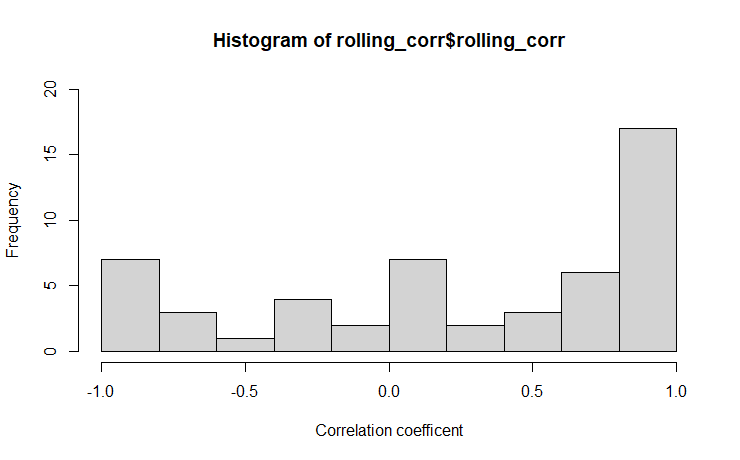

**Figure 3. Histogram of the frequency distribution of 3-day rolling Pearson correlation coefficients estimated using ONS and Virus Watch estimated COVID-19 incidence rates in Wales from 22 June 2020 to 13 June 2022.**

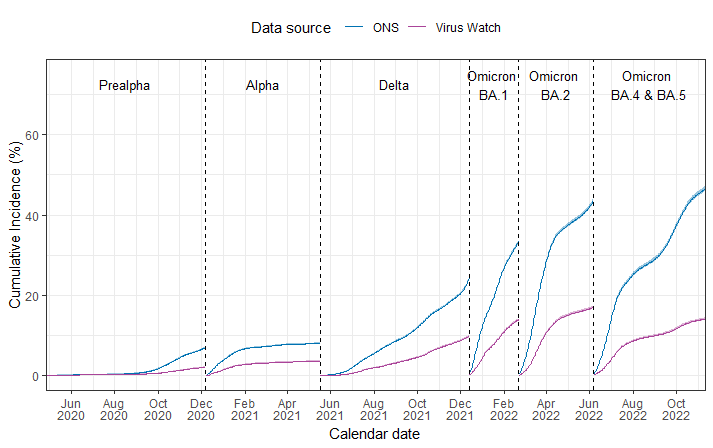

**Figure 4. Virus Watch and ONS estimated cumulative incidence by period (dominant variant of concern) in England from 26 April 2020 to 11 November 2022.**

**Table 1. Virus Watch and ONS estimated cumulative incidence by period (variant) for all ages in England from 26 April 2020 to 11 November 2022.**

|  | **Virus Watch**  Cumulative Incidence  (95% Confidence Interval) | **ONS**  Cumulative Incidence  (95% Credible Interval) |
| --- | --- | --- |
| pre-Alpha | 2.1% (2.0% - 2.3%) | 7.0% (6.9% - 7.2%) |
| Alpha | 3.6% (3.5% - 3.8%) | 8.1% (7.9% - 8.2%) |
| Delta | 9.8% (9.5% - 10.0%) | 24.2% (23.9% - 24.5%) |
| Omicron BA.1 | 14.0% (13.7% - 14.3%) | 33.6% (33.1% - 34.0%) |
| Omicron BA.2 | 17.0% (16.7% - 17.4%) | 43.6% (43.1% - 44.1%) |
| Omicron BA.4 & 5 | 14.1% (13.8% -14.5 %) | 46.5% (45.9% - 47.1%) |

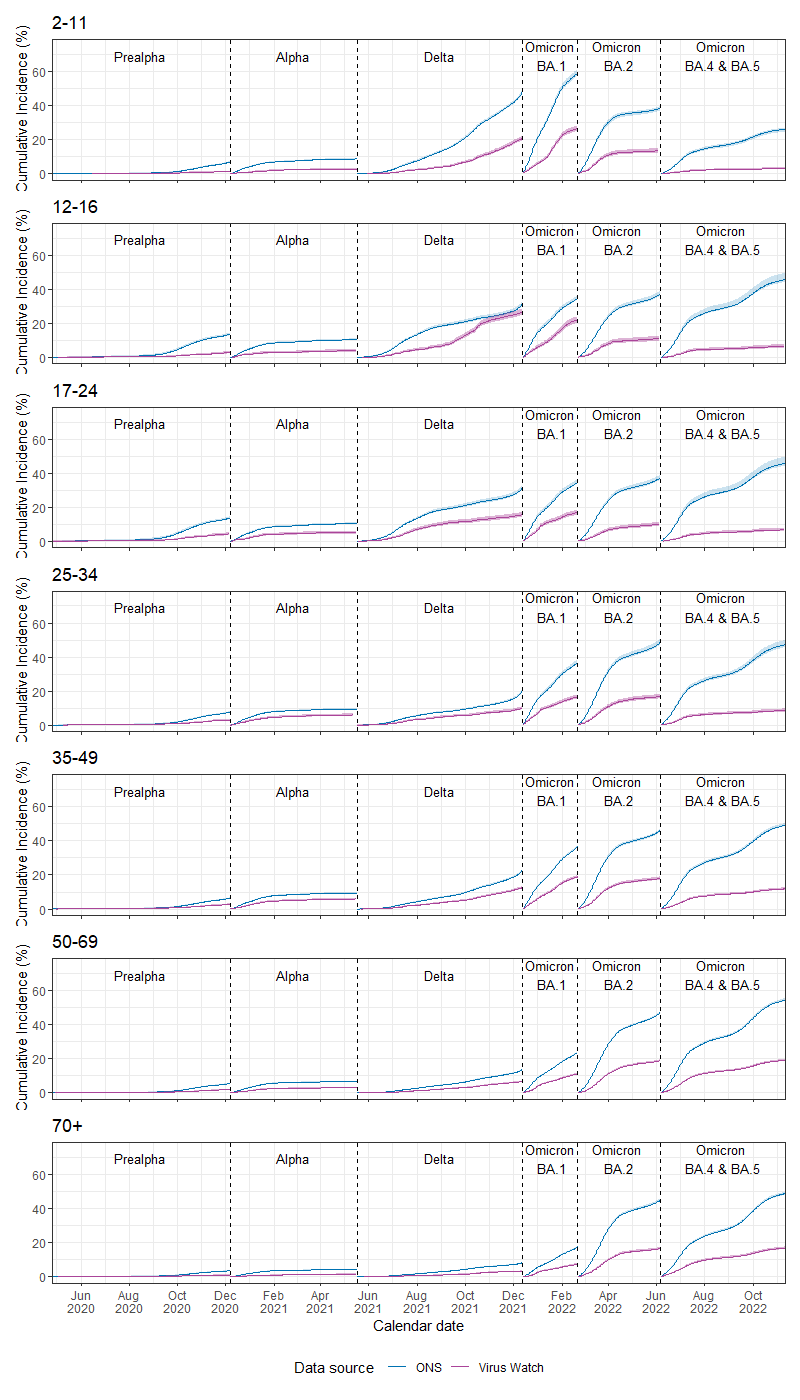

**Figure 5. Virus Watch and ONS-estimated cumulative incidence by period (dominant variant of concern) and age group in England from 26 April 2020 to 11 November 2022.**
